# Technology transfer of a dried blood virus neutralization assay to a GAVI-eligible country

**DOI:** 10.1101/2024.07.23.24310857

**Authors:** Evangeline Obodai, Jonne Terstappen, Jude Y. Mensah, Anouk Versnel, Comfort N. Antwi, Louis J. Bont, Daniela Cianci, Eveline M. Delemarre, John K. Odoom, Peter M. van de Ven, Natalie I. Mazur

**Affiliations:** Department of Virology, Noguchi Memorial Institute for Medical Research, University of Ghana, Legon, Accra, Ghana; Department of Pediatric Infectious Diseases and Immunology, Wilhelmina Children’s Hospital, University Medical Center Utrecht, Utrecht, The Netherlands; Center for Translational Immunology, University Medical Center Utrecht, Utrecht, The Netherlands; ReSViNET Foundation, Zeist, The Netherlands; Julius Center for Health Sciences and Primary Care, Department of Data Science & Biostatistics, University Medical Center Utrecht, Utrecht University, Utrecht, The Netherlands

## Abstract

**Background:** Global health clinical research is commonly led by high-income countries (HICs) as low- and middle-income countries face barriers to participate, including lack of financial and human capacity and lack of research environment. Respiratory syncytial virus (RSV) vaccine development is also led by HICs, preventing global vaccine access while LMICs carry the burden of life-threatening disease. This study aims to transfer an RSV neutralization assay, which uses live cells and virus with inherent high variation, to a GAVI-eligible country.

**Methods:** Using a train-the-trainer approach, a Ghanaian researcher was trained in the Netherlands on the dried blood-based RSV neutralization assay. Subsequently, a Dutch researcher visited Ghana to support the process of adapting the technique to the Ghanaian setting. In a previously validated RSV neutralization assay on dried blood, Hep-2 cells were infected with a serial dilution of sample-virus mixture to determine the half-maximal inhibitory concentration. Fifty-one dried blood and serum samples were tested in parallel in both countries to assess concordance.

**Results:** Training and technology transfer was deemed successful, which was defined as neutralization measurements by the Ghana team and high concordance (Lin’s concordance correlation coefficient (CCC) > 0.8). Neutralizing capacity measured in identical samples in Ghana and the Netherlands correlated highly (Lin’s CCC = 0.87; Spearman rho = 0.89) but was systematically lower in Ghana than the Netherlands.

**Conclusion:** We show successful strengthening of the laboratory research capacity in a GAVI-eligible country. Reliable measurement of RSV neutralizing antibodies in a GAVI-eligible country and the use of dried blood can contribute to inclusion of LMICs in RSV vaccine development and access.

**Funding:** None

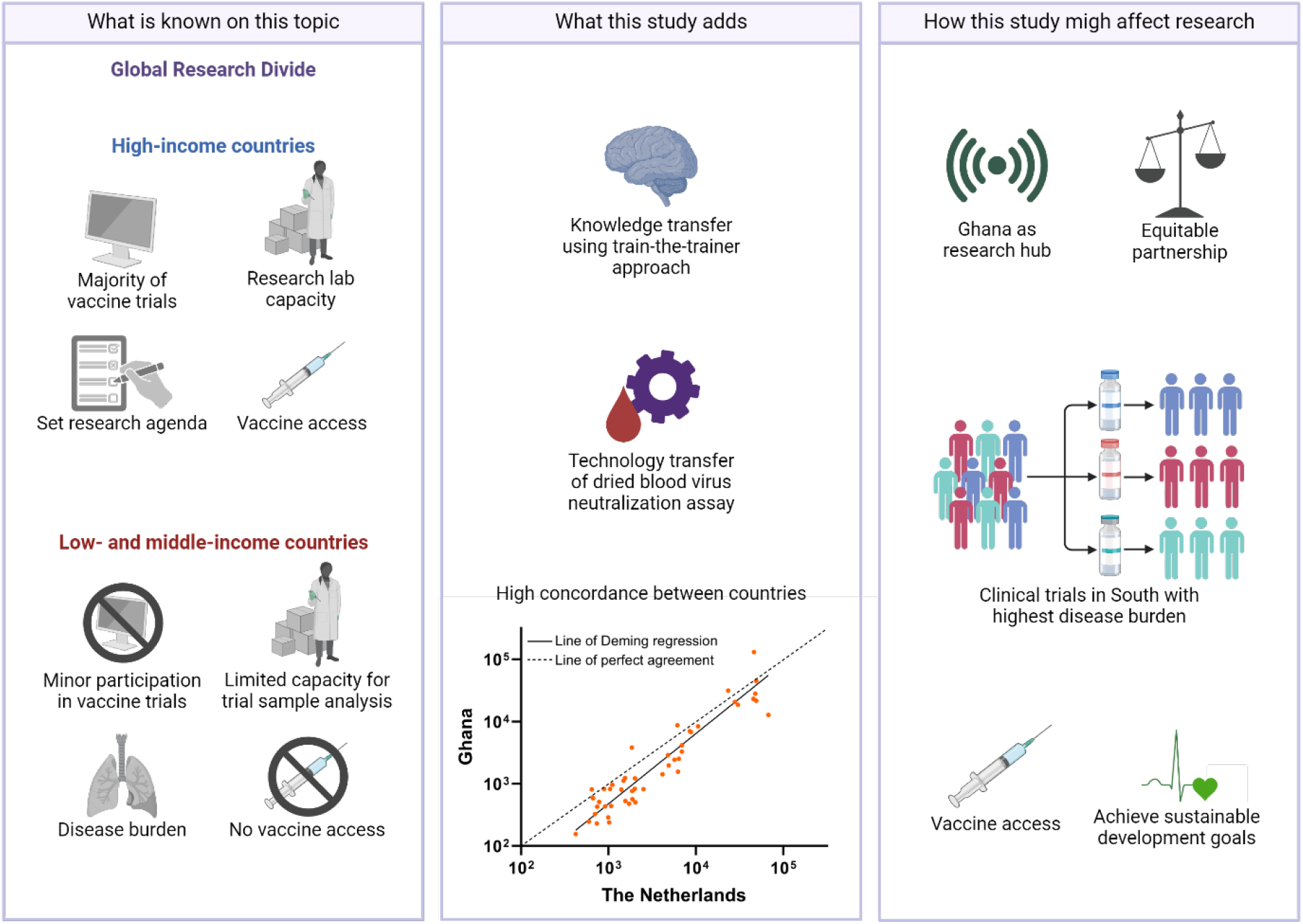

**KEY MESSAGES:** *What is already known?:* The global health research divide is evident in respiratory syncytial virus (RSV) vaccine development, where vaccines are developed and available in high-income countries (HICs), despite low- and middle-income countries (LMICs) bearing the majority of the disease burden. Representative trials and research capacity strengthening in LMICs are needed to ensure global vaccine access and equity.

*What this study adds?:* In this newly established partnership, we used a train-the-trainer approach to transfer a low-tech dried blood live-virus neutralization assay from the Netherlands to Ghana, a GAVI-eligible country. A sample panel measured in parallel in both countries showed high concordance and correlation despite the difficulties of running a bioassay with inherently high variability while using distinct materials.

*How this study might affect research, practice, or policy?:* The Ghana laboratory will leverage the established RSV neutralization assay to serve as a research hub in Africa supporting other LMICs to perform clinical sample analysis and management of clinical trial samples, ultimately reducing barriers to clinical trials and vaccine access, thereby advancing the sustainable development goals.

## INTRODUCTION

The global divide in resources, including research funding and scientific capacity, results in a research agenda set by high income countries (HICs).^1^ Low- and middle-income countries (LMICs) experience substantial barriers to participate in clinical research, including lack of funding, lack of skilled personnel, and lack of research environment.^2^ Equitable partnerships can facilitate research capacity strengthening by sharing knowledge, technology, expertise, and funding to effectively address pertinent research questions and enhance self-sufficiency.^1 3^

The global research divide is visible in the respiratory syncytial virus (RSV) vaccine development: vaccines and immunization are predominantly developed in HICs, where RSV is now a vaccine-preventable disease,^4-7^ although more than 95% of RSV-associated hospitalizations and deaths occur in LMICs.^8^ Registration trials included participants in LMICs but sample and data analysis were performed in HICs. Consequently, no RSV vaccine or monoclonal antibody (mAb) is within reach in LMICs despite the high burden.^9 10^ Representative clinical trials in LMICs are key to ensure global access and affordability of an effective RSV vaccine.^9 11^ Trials largely monitor neutralizing antibody (nAb) levels as a correlate of protection, however a simple tool to measure protection after RSV vaccination is lacking in LMICs due to prohibitive costs and lack of research capacity.^12^ Bioassays can measure nAbs with live cells and virus but the live components inherently increase the complexity and the variation in results. Cell-based assays are recommended to have a target coefficient of variation (CV) of < 50% with an average of 25%,^13^ whereas industry guidelines are stricter with CV < 20%.^14^ Capacity strengthening for vaccine trial research in GAVI-eligible countries, i.e. LMICs that qualify for support from the GAVI Vaccine Alliance to improve access to vaccines and immunization services, could accelerate access to RSV vaccines while promoting research equity.

Serum is generally considered the gold-standard sample to measure surrogates of vaccine-induced protective immunity.^15^ Dried blood spots (DBSs), offer attractive alternatives to serum antibody testing and have already been found to be appropriate tools for the antibody surveillance and/or diagnosis of other viral pathogens such as HCV and HIV.^16 17^ Finger prick DBS samples provide numerous advantages over serum, as they are cheaper, less invasive, and easy to (self-)collect in clinical and nonclinical settings. DBS do not require specialist equipment for processing and storage, have long-term stability, are readily transportable and hence may facilitate clinical trials in LMICs.^18 19^ Our recently validated neutralization assay using dried blood offers a patient-centric and financial solution to logistical barriers to immunization research in LMICs.^20^

Here, we describe a technology and knowledge transfer of a dried blood-based RSV neutralization assay as part of a new partnership to strengthen the research capacity in a GAVI-eligible country. The low-tech low-cost assay can be used to measure clinical trial endpoints in a low resource setting and can set a platform for future development of broader respiratory virus surveillance, vaccine impact, and correlate of protection studies in LMICs.

## METHODS

### Site selection and training

The Virology Department of the Noguchi Memorial Institute for Medical Research (NMIMR) situated in Accra, Ghana was selected as a site after a screening process involving ten sites within the international GOLD-III ICU network.^21^ Thorough inquiry ensured availability of the required facilities to conduct the assay: cell and viral culture systems, biosafety level 2 cabinets (including lamina air flow hoods), fluorescence plate reader with required filters, and available research staff experienced in cell culture and virus handling.

Using the train-the-trainer approach,^22^ a researcher from NMIMR visited the University Medical Center Utrecht (UMCU), The Netherlands, for one month to be trained on the dried blood-based RSV neutralization assay.^20^ Subsequently a Dutch expert researcher visited Ghana for one month to support the process of adapting the technique to the Ghanaian setting (Table 1). Successful training was defined as measurement of neutralizing capacity in dried blood using the neutralization assay by the Ghana team.

**Table 1.** Distinct materials between the two laboratories.

| **Distinct materials potentially increasing variation** | **Distinct materials unlikely increasing variation** | **Actions to reduce variation** |
| --- | --- | --- |
| Virus stocks cultured from the same master stock | Fluorescence plate-reader | Triplicates rather than duplicates |
| HEp-2 cell stocks | Single-channel pipettes | WHO international standard |
| Culture medium | Non-critical reagents, e.g. PBS | Verification of data analysis* |
| Multichannel with securely fitting tips | General laboratory equipment, e.g. rocker, centrifuges, heat block |  |

\*Raw data from Ghana was analysed in parallel by both Ghana and the Netherlands teams and compared to cross-validate the analysis methods.

### Sample panel preparation and handling

Individual samples for validation of the assay were obtained from healthy adults from the UMCU healthy donor service (medical ethical approval 07-125/O) to be measured in parallel in both laboratories. Whole blood was either left blank or spiked with anti-RSV mAbs palivizumab and RSM01 (Bill & Melinda Gates Medical Research Institute) at concentrations ranging 5-200 μg/mL. After 30 minutes on a rocker to mix, Mitra 20 μl volumetric absorptive microsampling tips (VAMS; Trajan Scientific, Neoteryx Clamshell, 20004) were touched to the whole blood sample surface to absorb the fixed volume and left to dry for 3 hours at room temperature.^20^ Dried blood was packaged in zip-lock bags containing desiccant to allow further drying. Blood of eight healthy donors provided five unique samples per donor for a total of 40 dried blood samples. Additionally, four archived dried blood samples from a previous study^20^, four in-house serum pools, and three commercially available serum pools (BEI Resources NR-4021, NR-4022, and NR4023) were added to the sample panel for a total of 51 samples. Samples were shipped to NMIMR on dry ice within five days and stored at -80°C until analysis. Prior to analysis on the neutralization assay, dried blood samples were thawed for 30 minutes before opening the air-tight bag to prevent condensation. Dried blood was rehydrated by incubating VAMS tips in 200 μL of PBS in Eppendorf tubes overnight at room temperature on a shaker at 300 rpm.

### Neutralization assay

Neutralizing capacity of the dried blood samples was tested using a neutralization assay described previously.^20^ Briefly, Hep-2 cells (ATCC CCL-23) were seeded at a concentration of 0.5 × 10^6^ cells/well in 384-well black optical bottom plates (Thermo Scientific, 142761) and incubated for > 4 hours. Dried blood eluate and serum controls, including WHO 1^st^ International Standard for Antiserum to RSV (NIBSC code:16/284),^23^ were heat-inactivated at 56°C for 30 minutes. Dried blood eluate was then centrifuged at 15.000g for 10 minutes to remove cell debris. Serum and dried blood eluate (1:10 during elution) were serially diluted in 3-fold from starting dilution 1:5 or 1:2 (total dilution 1:20), respectively, in Dulbecco’s modified Eagle’s Medium containing 10% FCS. Recombinant mKate-RSV-A2^24 25^ was added 1:1 to the sample dilution series and incubated for 1 hour at 37°C before addition of 50 μL of the sample-virus mixture to the adherent Hep-2 cells. After 48 hours incubation at 37°C, relative fluorescence units were recorded using excitation at 584 nm and emission at 620 nm (FLUOstar OMEGA, BMG Labtech and Tecan SparkControl™ V3.2). The neutralization curve and 50% inhibitory dilution (ID50) of samples were analysed using a log(inhibitor) vs response 4-parameter nonlinear regression curve in GraphPad Prism version 8.3 or higher (GraphPad Software Inc., San Diego, CA, USA). International units (IU) were calculated according to manufacturer’s instruction.^23^ The bioassay using live cells and virus is inherently subject to high variability compared to non-biological assays, e.g. ELISA, with <50% variance considered as the target precision in cell-based assays.^13^ Healthy adults were expected to have pre-existing anti-RSV antibodies detectable in the assay at baseline (ID50 of approximately 10^3^) with an increase post-spiking with palivizumab or RSM01.^26^

### Sample size and statistical analysis

Results reported as IU/mL from both laboratories were compared using the Lin’s concordance correlation coefficient (CCC) to test how well the pairs of observations conform relative to perfect concordance using DescTools package in R (version 4.4.1).^27^ In case the data do not follow (near-to) normal distribution, Deming regression is performed using SimplyAgree package in R additional to Lin’s CCC. Deming regression takes into account the measurement errors in both the dependent and independent variables simultaneously.^28^ Additionally, Bland-Altman plots using the Log10 transformed data are used to describe the agreement between the two measurements.^29 30^ A small bias is expected between the two laboratories due the live biological components of the assay resulting in a difference unequal to zero. Simulations performed in R using precision data previously obtained at the UMCU showed that a sample size of 50 samples would provide acceptable Lin’s CCC values (≥0.9) and precision (confidence interval width ≤ 0.1).^27^ Successful technology transfer was defined as Lin’s CCC > 0.8. Assay precision was calculated as the percentage %CV of relative fluorescence units of sample duplicates (intra-assay precision) or %CV of the linear ID50 between runs (inter-assay precision). Plots were created in GraphPad Prism version 10.1.2.

### Patient involvement

Patients were not involved in the design or conduct of the study. Interim study results were presented to the ReSViNET patient advisory board.

## RESULTS

Successful training was achieved within the planned time frame of one month of remote training of the trainer and one month on-site training of other personnel, according to the pre-specified definition of measuring neutralizing capacity in dried blood using the neutralization assay by the Ghana team. The RSV neutralization assay performed in Ghana and the Netherlands detected neutralizing activity in all blood samples of the sample panel including blank dried blood samples. Two samples gave unreliable ID50 values according to quality control criteria^20^ upon repeated measurements in Ghana and were therefore excluded from analysis. Lin’s CCC was 0.87 (95% confidence interval (CI) 0.80-0.92), which is above the defined value of 0.80 for a successful transfer (Figure 1). As the data were minimally skewed, we also performed Deming regression resulting in the following equation: Ghana [IU/mL] = 1.13 x Netherlands – 2.37 (95% CI of slope 0.9795 to 1.280, 95% CI of Y-intercept -4.057 to -0.6775). Values in Ghana highly correlated with those in the Netherlands over the range of ID50 values (Spearman rho = 0.89, 95% CI 0.81-0.94, p<0.0001) but were systematically lower. The measurement differences in RSV neutralizing capacity in dried blood have a significant bias, better illustrated in the Bland-Altman plots (Figure 2). The Log10 mean difference (Netherlands – Ghana) was 0.26 (Figure 2), corresponding to a geometric mean difference of 1.81 with more bias seen in samples with lower neutralizing capacity. Inter-assay and intra-assay precision were lower in Ghana than in The Netherlands (Table 2).

**Table 2.** Assay precision in the two laboratories.

|  | Ghana | The Netherlands |
| --- | --- | --- |
| Inter-assay precision (median, IQR) | 35.6 (30.5 - 37.5) | 23.6 (22.5 - 26.4) |
| Intra-assay precision (median, IQR) | 12.9 (5.1 – 26.2) | 3.5 (1.6 - 6.6) |

Precision expressed as the %coefficient of variation (%CV) where lower %CV equals higher precision. IQR, interquartile range.

**Figure 1.**
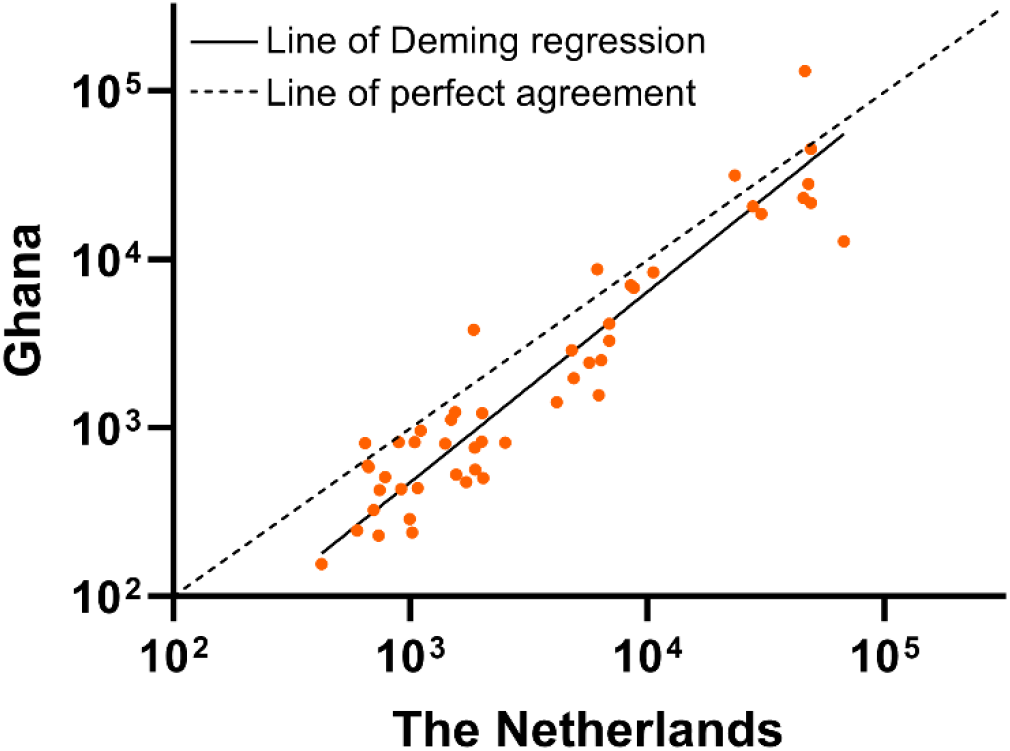
Scatter plot of RSV neutralizing capacity in blood samples measurements in Ghana over measurements in the Netherlands. Scatter plot for Log_10_-Log_10_ transformed RSV neutralizing antibodies (ID50 in IU/mL) in dried blood measured in Ghana over measurements in the Netherlands (n=49 paired measurements). Deming regression (solid line) is compared to the line of perfect agreement (dashed line) to calculate the Lin’s concordance correlation coefficient of 0.87. ID50, dilution factor with 50% viral neutralization activity; IU/mL, international units per milliliter.

**Figure 2.**
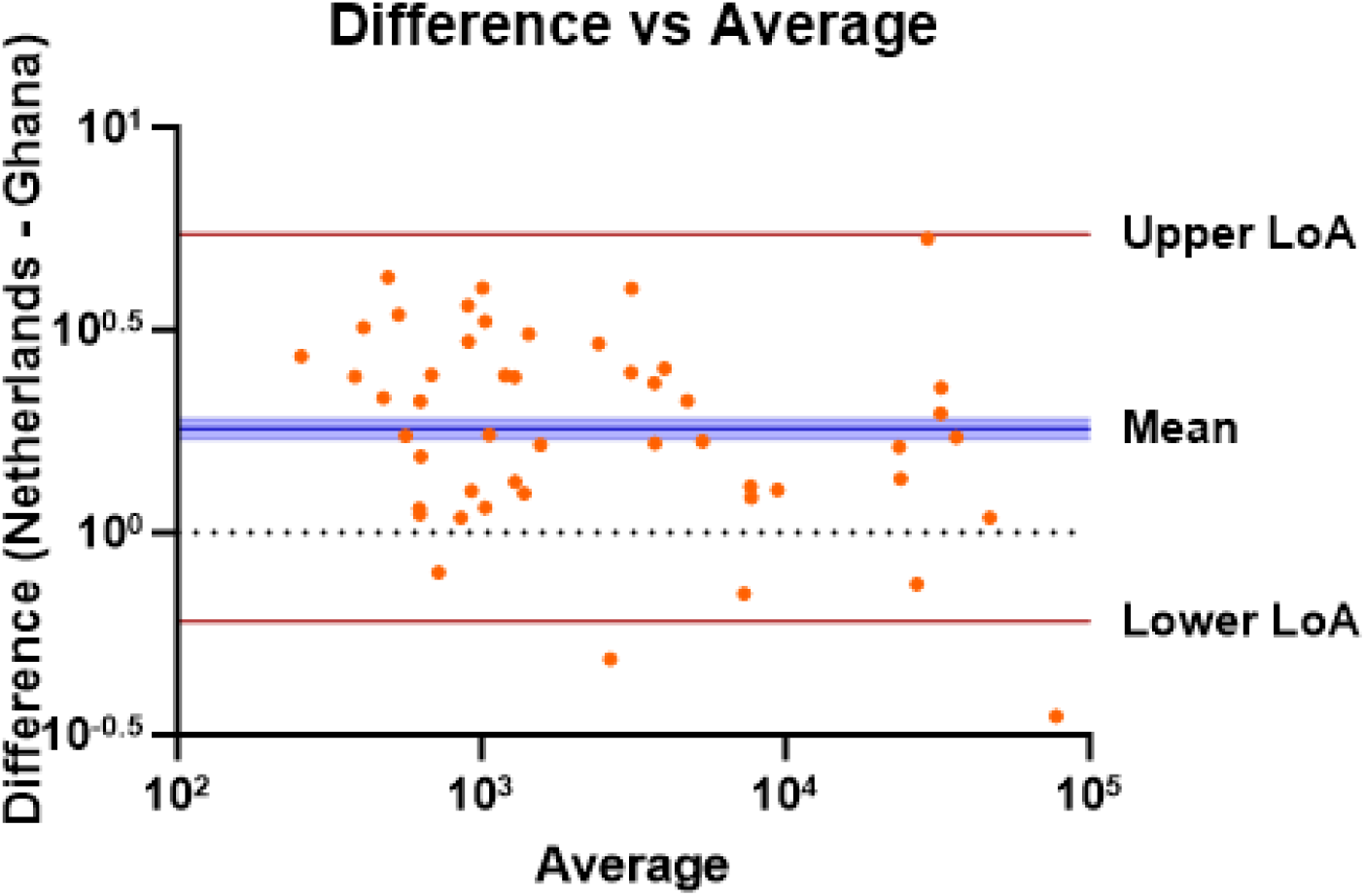
Bland-Altman plot of the agreement for RSV neutralizing antibody measurements between the Netherlands and Ghana. Scatter plot of difference between Log_10_ transformed neutralizing antibodies (ID50 in IU/mL) in dried blood samples measured the Netherlands and Ghana vs. the average of the two measurements. The mean difference (bias) with 95% CI in blue. Limits of agreement are calculated as mean bias ± 1.96*standard deviation. CI, confidence interval; ID50, dilution factor with 50% viral neutralization activity; LoA, limit of agreement.

## DISCUSSION

We show successful strengthening of the laboratory research capacity in Ghana by implementing a virus neutralization assay with dried blood. The neutralizing antibodies in matched dried blood samples obtained at the laboratories in Ghana and the Netherlands had high concordance (Lin’s CCC = 0.87) and were highly correlated (Spearman rho = 0.89) in the context of a biological assay with high expected variance due to the use of live virus and live cells. The reliable measurement of neutralizing antibodies acts as proof-of-principle that knowledge and technology can be successfully transferred and thereby promote the participation of LMICs in RSV clinical trials.

The four recently available RSV vaccines and mAbs are only in use in HICs, despite the high burden in LMICs.^9^ Urgent measures are needed to minimize the existing vaccine gap, including conducting representative trials and reducing pricing.^9 10^ Notably, all phase 3 registration trials of the recently market approved RSV vaccines were recruiting participants in upper middle-income countries (UMICs)^5-7 31^ yet these participants still lack post-trial access to these vaccines and no low or lower-middle income countries participate in the trials, except for The Gambia. To our knowledge, all trial samples are shipped to HICs for neutralizing antibody measurements, further exposing the disparities and limited roles left for LMICs in addressing global health challenges posed by RSV. During the COVID-19 pandemic, LMICs played a crucial role in the genetic surveillance identifying mutants of concern.^32-34^ However, vaccines developed in HICs tend to induce lower immune responses in LMICs,^35^ underscoring the importance to conduct vaccine trials in LMICs. The research community shares the responsibility to include LMICs in global health research efforts, for example by establishing partnerships.^1^ The current assay set-up in Ghana, a GAVI-eligible country, and use of dried blood could facilitate clinical trials in LMICs, thereby ensuring access to effective vaccines and mAbs, as well as addressing other research priorities within the local research agenda. Dried blood sampling, more accepted than serum due to its common use in HIV testing,^17^ is also more practical and safer for self-sampling or acquisition by field technicians and does not require cold-chain transportation, making it more suitable for LMIC settings. However, the lack of local production of VAMS devices and the price could pose a challenge in the procurement of VAMS for large-scale trials.

The current study had several strengths and limitations. The on-site training of the leading Ghanaian investigator in the Netherlands allowed an active practical hands-on experience and collaborative knowledge sharing. These fostered real time feedback, troubleshooting, seamless transfer of expertise between the two labs within a short time and contributed to the overall success of the tech transfer. By training the trainer, consistency in methodology and peer-to-peer learning was obtained in all Ghana trained personnel. Moreover, the use of the WHO international standard facilitated direct comparison of the assay results. Although identical methodology was used, certain critical reagents such as growth medium, virus, and cell culture, varied between the laboratories. The variations observed in the data were partly corrected by normalizing the ID50 values to IUs with the international standard. In industry, it is commonly considered best practice to acquire identical equipment and materials in the context of a technology transfer, which was not feasible in the current study. The lower resource setting in Ghana led to discrepancy in the availability of materials, such as a multichannel with securely fitting pipette tips (Table 1). Inter-assay precision was higher in the Netherlands than in Ghana potentially due to higher quality of materials (Table 2). Despite these limitations of using varying materials of varying quality with identical methodology, we were able to obtain a high correlation and concordance. A third limitation was the lack of cross-validation from Ghanaian samples shipped to the Netherlands, which is planned in a future study. The shipment of samples and consequently later measurement of samples in Ghana potentially caused the systematically lower neutralizing capacity in Ghana than in the Netherlands. As there was no thawing during the shipment and antibodies are generally stable, we expect limited impact of the lack of cross-validation. Fourth, we transferred a neutralization assay using RSV-A that cannot measure RSV-B neutralizing antibodies. Assessment of protection against both RSV subtypes would require adaptation of the assay. Lastly, we observed a bias in measurements between the labs with higher measurement bias in samples with lower neutralization capacity. As the number of observations is small for higher averages, the variation in bias could potentially be an artefact of lack of normal distribution in ID50 values. Cell-based bioassays typically target a variance below 50%, averaging around 25%,^13^ so the observed 81% mean bias is likely partially attributable and within acceptable assay variation. As the neutralization is best used to measure relative nAb differences in clinical trials, the lack of full agreement might be acceptable.

The current research project has significantly impacted RSV clinical trial capacity in Ghana, establishing the laboratory as a research hub supporting other African countries. By successfully transferring the dried blood-based RSV neutralization assay technology to Ghana, local researchers are now equipped and planning to analyze RSV clinical trial samples. Taking away some of the barriers to active participation of LMICs, especially GAVI-eligible countries, in clinical trials improves the access to vaccines where the disease burden is highest. This capacity strengthening not only enhances Ghana’s role in regional health research but also opens opportunities for expanding similar capabilities to labs in other regions and countries. Importantly, establishment of the neutralization assay empowers Ghanaian scientists to participate in and lead research relevant to the population, ensuring that local health challenges are addressed with local expertise. This project contributes to decolonizing global health by shifting the balance of research capabilities and decision-making power from HICs to LMICs through equitable partnerships with team engagement at all stages of the study. Consequently, it fosters a more equitable global health landscape where research and innovation are driven by those most affected by the diseases studied in order to achieve the 2030 sustainable development goals.^1^

## CONCLUSION

We provide a proof-of-principle that a bioassay with inherently high variability can be transferred to resource limited settings such as in Ghana with limited time and financial investment. Strengthening research-capacity in LMICs offers a platform for future development of a broader vaccine development, vaccine impact, and correlate of protection studies in LMICs. The transfer of this assay is a key step in minimizing the RSV vaccine gap between HICs and LMICs.

## Acknowledgements

We thank Tracy T. Ruckwardt and Alexandrine Derrien-Colemyn from the National Institute of Health, USA, for the material and methodological support. We thank Joleen T. White from the Bill & Melinda Gates Medical Research Institute for providing RSM01. We thank the Center of Translational Immunology for the support during training. This work was not supported by external funding.

## Contributors

All authors were involved in the review and editing of the work. EO and JT contributed to methodology, investigation, resources, data curation, validation, project administration, and writing of the original draft. JYM, AV, and CNA contributed to investigation and data curation. DC designed the statistical methodology and PvdV performed the formal analysis. LJB, EMD, JKO, and NIM supervised the study. LJB, NIM, EO and JT conceptualized the project.

## Data availability

On reasonable request to the corresponding authors, the data can be shared with publication.

